# Higher mortality in men from COVID19 infection-understanding the factors that drive the differences between the biological sexes

**DOI:** 10.1101/2020.04.19.20062174

**Authors:** Swaminathan P. Iyer, Joe Ensor, Kartik Anand, Patrick Hwu, Vivek Subbiah, Christopher Flowers, Rudragouda Channappanavar

## Abstract

The emergent global pandemic caused by the rapid spread of Severe Acute Respiratory syndrome-Coronavirus-2 (SARS-CoV-2) has led to increased mortality and negatively impacted day to day activities of humankind within a short period of time. As the data is rapidly emerging from earlier outbreak locations around the world, there are efforts to assimilate this with the knowledge from prior epidemics and find rapid solutions for this. One of the observations and a recurring theme is the disproportionate differences in the incidence of infection and the consequent mortality between males and females. We, therefore, analyzed retrospective datasets from the previous epidemics and the ongoing pandemic in order to address these differences in clinical outcomes. The data shows that even though the infection rates are similar, the odds ratio of male mortality remains high, indicating a divergence in the crosstalk between the three pathogenic human Coronavirus (hCoVs)-the SARS-CoV, MERS-CoV and the SARS-CoV-2 and immune effectors in the two sexes. One proximate cause is the sex-specific modulation of the X-linked genes that can influence susceptibility to infection. Future studies are needed to confirm these findings, which can form the basis for developing rational strategies for ending the current and preventing future pandemics.

## Introduction

The World Health Organization (WHO) has recently declared SARS-CoV-2 initiated **<u>Co</u>**rona**<u>vi</u>**rus **d**isease 20**19** (COVID19) as a pandemic^1,2^. SARS-CoV-2 belongs to the coronaviruses (hCoVs) family that also include strains such as SARS-CoV, MERS-CoV, OC43, 229E, NL63, and HKU1^3^. The latter four are generally associated with self-limiting respiratory tract infections. However, unlike the seasonal strains, the SARS-CoV, MERS-CoV, and SARS-CoV2 strains cause outbreaks of severe respiratory disease in humans^4-10^. While SARS- and MERS-CoV spread among humans was limited, the current strain of SARS-coV-2 is rapidly being transmitted from human to human by aerosol or by direct contact leadings to severe symptoms with accompanying pneumonia and acute respiratory distress in many patients. There are many similarities between COVID 19 and previous two similar outbreaks of human CoVs (hCoVs)-SARS that occurred in 2003 and the Middle Eastern Respiratory Syndrome (MERS) in 2012, in terms of the epidemiology, immunology and the extreme clinical manifestations. The two epidemiologic parameters include the basic reproduction number (R0) and the best estimates of case-fatality rates (CFR)*^11^. The global epidemic of the 2002-2003 SARS outbreak caused over 8000 infections, including 774 deaths in 29 countries and an estimated CFR of 9.6% globally, while the MERS outbreak was estimated to have a much higher CFR of 34.5% with more than 2500 infections to date^12^. Although several studies have shown an estimated R0 of 2.2 for COVID19, there is a broad range of values for the CFR given the inability to estimate during the early phase of this outbreak realiably. The risk for mortality shows an increase in the elderly population^13^ and in those with certain pre-existing conditions related to decreased or marginal cardiac, pulmonary, renal functions, and immunosuppressive states. Another area that is gaining attention is the biological sex differences in the incidence of infection and CFR*

## Methods

Studies chosen for these analyses were based on the availability of the sample size, and that of detailed patient demographics, including gender-associated incidence and mortality in peer-reviewed journals. The COVID19 sample set included the first three largest retrospective reports from China as they describe the unfolding clinical events and hospitals were trying to grapple with this sudden outbreak of the virus. Chen et al. describe a retrospective study from Wuhan Jinyintan hospital in the first three weeks of January. By that time, testing was available, and Guan et al. describe another retrospective case series at the First Affiliated Hospital of Guangzhou Medical University. Wang et al. describe the clinical outcomes of 138 patients with SARS-CoV-2 infected pneumonia at the Zhongnan Hospital of Wuhan University in Wuhan, China, between January 1 to January 28, 2020, with the final date of follow-up on February 3, 2020. For the SARS-CoV in 2003, the two studies include those compiled by Booth et al. in Toronto and another larger series by Karlberg et al. from China, Hong Kong, Special Administrative Region (Hong Kong SAR). The data set from the latter was derived from the published WHO report in 2003. Finally, the dataset for MERS-CoV includes the two largest series by Chen et al. and Alghamdi et al. that have detailed comparative epidemiology covering Saudi Arabia and South Korea. In order to accommodate statistical heterogeneity between these retrospective studies, we utilized the random effects model that assumes that different studies have different true exposure effects^14^. For individual studies, incidence and death rates are presented as percentages with 95% Wilson score confidence intervals. The percentage of male cases was compared to 50% using a one-sample binomial test for proportions. Within the study, death rates were compared between males and females using a chi-square test. Results were aggregated across studies, and gender differences were compared using the GLIMMIX procedure of SAS to fit random effects model due to heterogeneity present in the datasets. All analyses were conducted using SAS 9.4, and statistical significance was defined as p < 0.05^15^.

## Results

### Biological sex differences between males and females in Incidence and mortality

Sex-specific differences in disease incidence and severity were seen during the previous pathogenic hCoV outbreaks^7-10^. The incidence of COVID-19 and MERS seems to be more common among males, but SARS appears to be more common among females (Table 1 and Table 3a). The SARS epidemic in 2003 showed differences in incidence and mortality between males and females. Out of a total of 8098 SARS infections from 27 different countries-slightly more than half were female (World Health Organization, 2003). However, females had lower mortality rates from SARS than males, a pattern that is maintained after adjusting for age^16^. In the data reported from Hong Kong SAR, the mortality rate for males was 21.9%, which was substantially higher than the mortality rate for females of 13.2%^10^. The table 1 below is based on three studies from China^4,5^. If one investigates whether the probability of being male is equivalent to being female (i.e., 50%), one would conclude that males were more likely to have the infection. Further review of CFR from SARS, MERS, and the ongoing COVID indicate a preferential outcome according to biological sex (Table 2 and Table 3b). In terms of mortality, males had increased deaths compared to females from MERS, SARS-CoV, and SARS-CoV2. (Table 2). The odds ratio of male mortality was 1.51 95%CI (1.08-2.08) in a study by Chen et al. looking at cases from Saudi Arabia and South Korea^7^. The odds ratio of male mortality was higher at 2.2 95% CI (1.37-3.51) in the study of cases just from Saudi Arabia alone (Table 2). The odds ratio of male mortality was also higher, at 1.85-95% CI (1.43-2.3) from SARS-CoV^9,10^. For the ongoing COVID19, the mortality data being collected is not entirely categorized by biological sex for some countries represented in Global Health 5050 COVID-19 data tracker^17^(Table 3b). Assuming a random effects model, the Global Health data yields an aggregate odds ratio (OR) of 1.84 for comparing the odds of death for males to females. There is statistically significant evidence in these studies strongly hinting at possible sex differences in the immune response.

**Table 1:**
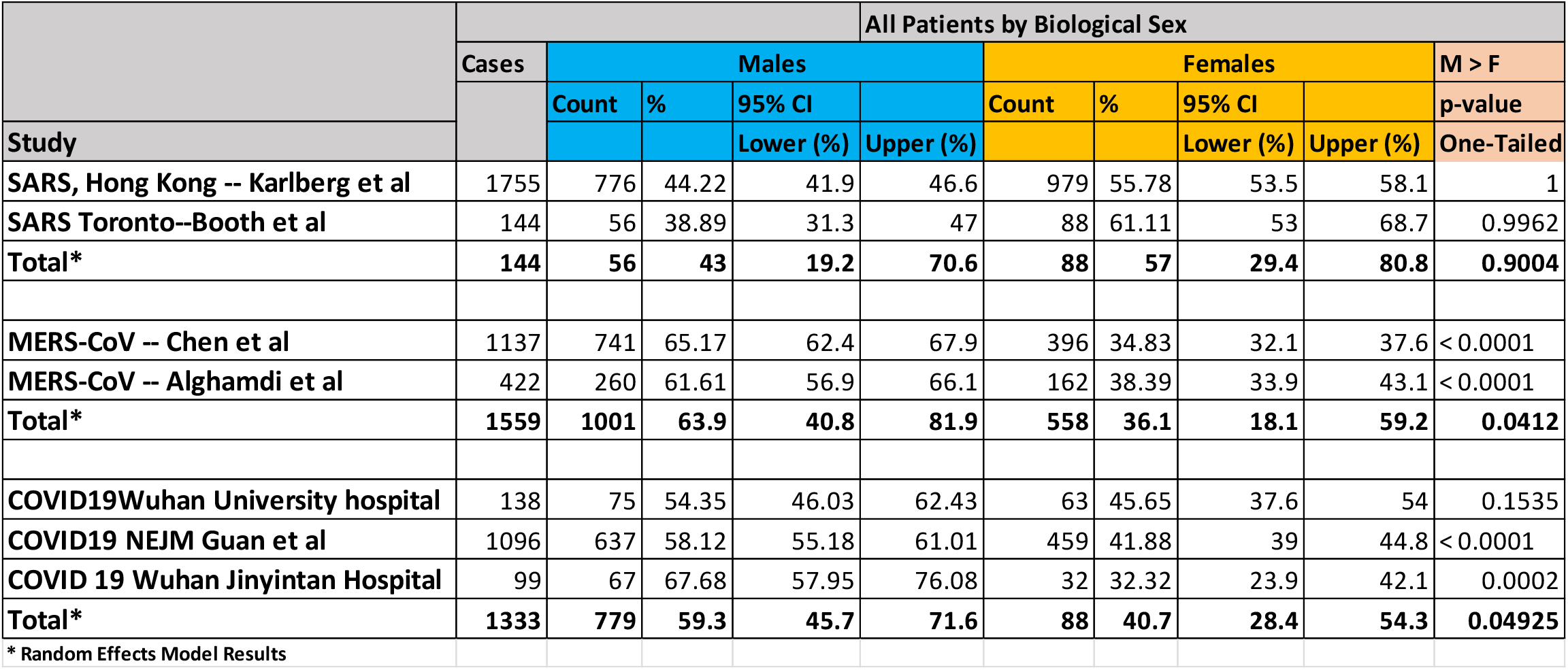
All Patients by Biological sex- incidence.

**Table 2:**
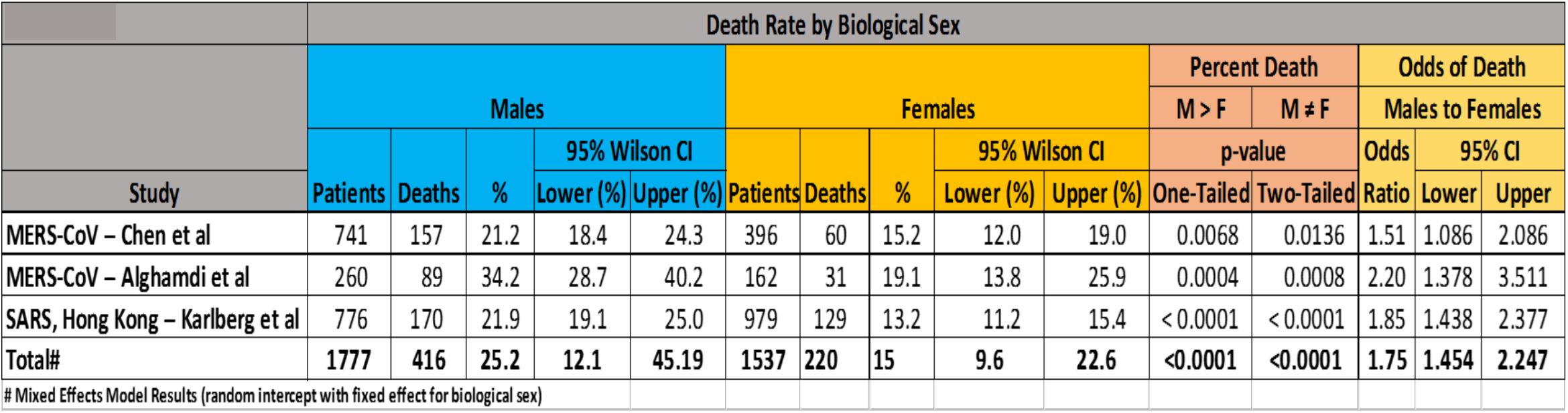
Death Rate by biological sex for SARS and MERS.

**Table 3:** COVID19 All Patients by Biological sex-incidence.

| Table 3 |  | COVID19 All Patients by Biological Sex- Incidence |  |  |  |  |  |  |  |
| --- | --- | --- | --- | --- | --- | --- | --- | --- | --- |
|  |  | Males |  |  | Females |  |  | M > F | M ≠ F |
|  |  | 95% Score CI |  |  | 95% CI |  |  | p-value |  |
| Country | Cases | % | Lower | Upper | % | Lower | Upper | One-Tailed | Two-Tailed |
| Italy | 57,989 | 58 | 57.6 | 58.4 | 42 | 41.6 | 42.4 | < 0.0001 | < 0.0001 |
| China | 55,924 | 51 | 50.6 | 51.4 | 49 | 48.6 | 49.4 | < 0.0001 | < 0.0001 |
| Germany | 36,508 | 55 | 54.5 | 55.5 | 45 | 44.5 | 45.5 | < 0.0001 | < 0.0001 |
| Spain | 25,113 | 51 | 50.4 | 51.6 | 49 | 48.4 | 49.6 | 0.0008 | 0.0015 |
| Iran | 14,991 | 57 | 56.2 | 57.8 | 43 | 42.2 | 43.8 | < 0.0001 | < 0.0001 |
| Switzerland | 12,161 | 49 | 48.1 | 49.9 | 51 | 50.1 | 51.9 | 0.9862 | 0.0276 |
| South Korea | 9,332 | 39 | 38.0 | 40.0 | 61 | 60.0 | 62.0 | 1.0000 | < 0.0001 |
| The Netherlands | 8,603 | 50 | 48.9 | 51.1 | 50 | 48.9 | 51.1 | 0.5000 | 1.0000 |
| Austria | 7,557 | 53 | 51.9 | 54.1 | 47 | 45.9 | 48.1 | < 0.0001 | < 0.0001 |
| Belgium | 7,284 | 50 | 48.8 | 51.2 | 50 | 48.8 | 51.2 | 0.5000 | 1.0000 |
| France | 6,378 | 47 | 45.8 | 48.2 | 53 | 51.8 | 54.2 | 1.0000 | < 0.0001 |
| Norway | 3,581 | 52 | 50.4 | 53.6 | 48 | 46.4 | 49.6 | 0.0084 | 0.0169 |
| Sweden | 3,046 | 53 | 51.2 | 54.8 | 47 | 45.2 | 48.8 | 0.0005 | 0.0010 |
| Portugal | 4,268 | 46 | 44.5 | 47.5 | 54 | 52.5 | 55.5 | 1.0000 | < 0.0001 |
| Canada | 4,043 | 52 | 50.5 | 53.5 | 48 | 46.5 | 49.6 | 0.0057 | 0.0113 |
| Australia | 3,010 | 52 | 50.2 | 53.8 | 48 | 46.2 | 49.8 | 0.0144 | 0.0287 |
| Denmark | 1,715 | 60 | 57.7 | 62.3 | 40 | 37.7 | 42.3 | < 0.0001 | < 0.0001 |
| **Total"** | **261,503** | **51.5** | **48.9** | **54.0** | **48.5** | **46.0** | **51.1** | **0.1195** | **0.2389** |
| " Random Effects Model Results |  |  |  |  |  |  |  |  |  |

**Table 4:**
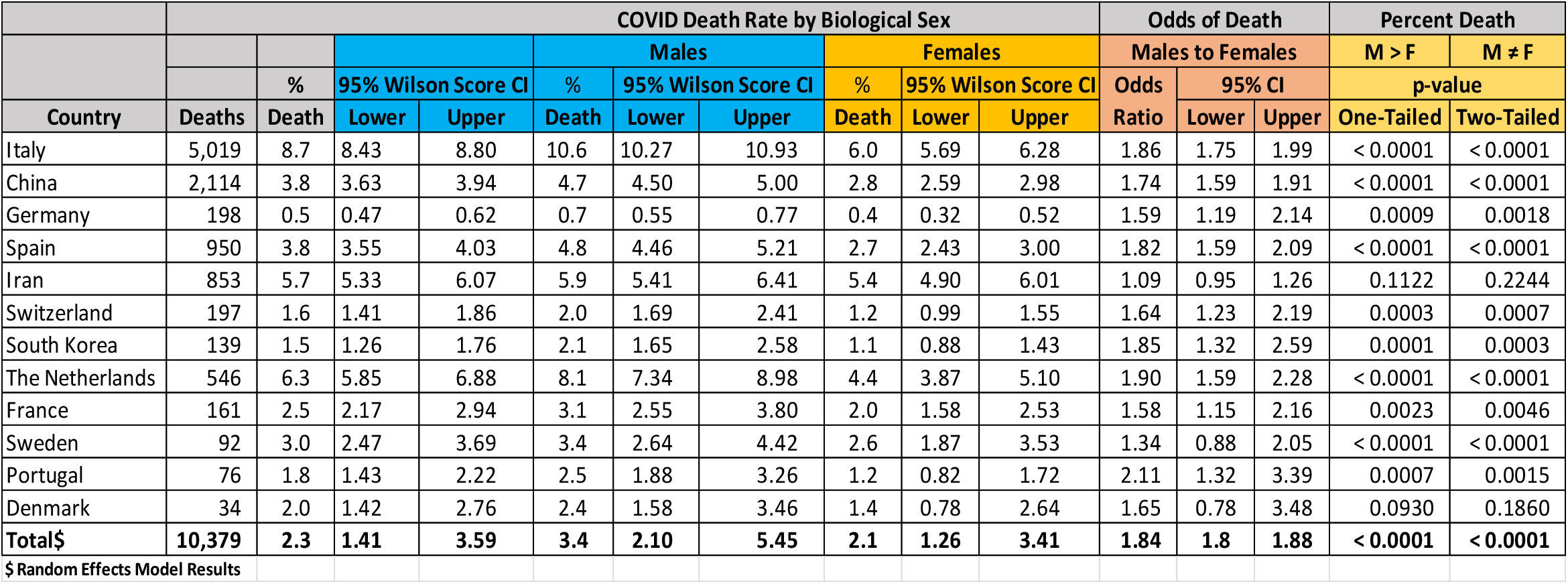
COVID19 Death Rate by Biological sex.

## Discussion

There are reported differences in the rate of infection and mortality rates between men and women. Disaggregated data collections are necessary to understand the underlying biological and socio-economic (including behavioral) factors causing this. We have shown empiric data that the mortality rate for males is higher and statistically significant compared to females from the three hCoV outbreaks of MERS, SARS-CoV, and SARS-CoV2^4-10^. The data across show that there were 8-16 additional men with the disease for every 100 women infected. Guan et al. study showed higher male numbers, with 58% of 1099 reported. In the US, among 1,482 COVID19 patients reported from COVID-19–Associated Hospitalization Surveillance Network (COVID-NET), 74.5% were aged ≥50 years, and 54.4% were male^18^. It is also clear that men made a significant proportion of certain risk high-risk groups such as the elderly population and in those with certain pre-existing conditions related to decreased or marginal cardiac, pulmonary, renal functions, and immunosuppressive states continue to remain vulnerable. Data from the China CDC indicate that the large numbers of deaths were elderly Chinese men with CFR of 14.8% in people 80 or older as opposed to 1.3% in 50-year old cohort and <0.5% in the younger population. The likely attributes here include a significant number of preexisting illnesses in men related to smoking and cardiovascular disease. In the COVID-NET dataset, the hospitalization rate was highest (13.8) among adults aged ≥65 years. Among 178 adult patients, 89.3% had one or more underlying conditions, including hypertension (49.7%), obesity (48.3%), chronic lung disease (34.6%), diabetes mellitus (28.3%), and cardiovascular disease (27.8%).

There is also a possibility that the apparent sex imbalance could reflect more exposure to the males because of frequent travel and contacts, as was reported in the MERS Saudi Arabian cohort. Since the virus appears to be transmitted primarily through large droplets and remains in aerosols for up to 3 days^19^, another factor that needs discussion is the gender differences in handwashing, particularly in public restrooms and airports. In a 2003 study of 175 individuals (95 women and 80 men) handwashing after restrooms use, indicated that women washed their hands with soap more often than men (61%vs. 37%) in the absence of a prompt to do so^20^. When a visual reminder was placed, the numbers went up for women but remained more or less the same for men (97% vs. 35%). Indeed emphasis on handwashing with soap and the extent of time is one of the most effective interventions to prevent the spread across all age-groups and gender^21^.

Other possibilities include fundamental biological factors that may provide better immune protection for women, which may be related to the estrogen and X-linked genes and X chromosome inactivation (XCI)^22^. XCI or Lyonization is the process through which one X chromosome is inactivated to balance the dosage of gene expression between XX females and XY males-this makes haploid males more susceptible to effects of the genetic variants. The X chromosome contains many genes that regulate the innate and adaptive immune system. These sex-based immunological differences have been shown to cause variations in susceptibility to infectious diseases and responses to vaccines and in the incidence of autoimmune diseases between males and females^22-24^. Some of these include pattern recognition receptors (PRRs) such as toll-like receptor (TLR)-TLR7 and TLR8, as well as Interleukin-1 receptor-associated kinase 1 (IRAK1), a key regulatory molecule in the TLR signaling pathway^23,24^. Preclinical animal studies have also validated this sexual dimorphism in the severity of pneumonia caused by various respiratory pathogens with the highest correlation for the innate immune response triggered at the early phase of infection^25^. Interestingly, Angiotensin converting enzyme 2 (ACE2), the receptor for both the SARS-CoV and the SARS-CoV-2 is encoded by its gene on Xp22. It has been shown that infection of human airway epithelia by hCoVs correlates with the state of bronchial epithelial cell differentiation and ACE2 expression and localization^26^. There is evidence of adipose tissue ACE2 being regulated by estrogens, contributing to sex differences in the development of obesity-related hypertension in rat models^27^. However, it is not clear if ACE2 genes are modulated differentially for SARS-CoV2 infection, although in a pre-print version on BioRxiv, Zhao et.al, show higher single-cell RNA expression profiling of ACE2 in two male donors compared to other 6 female donors^28^. Finally, another candidate gene is the X-linked glucose-6-phosphate dehydrogenase (G6PD); the deficiency of the protein due to mutations is the most common enzymopathy, manifesting as hemolysis due to oxidative stress. Given the high prevalence of this mutation in African-Americans (1 in 10), Mediterranean and Asians, and the predilection for increased hCoV infections in preclinical G6PD knockdown models, it is essential to elucidate its role in the outcomes to COVID infection^29,30^. In summary, the evidence currently available for X chromosome studies involving X-linked innate immune genes remain incomplete and needs additional investigations to see if they are driving the differences.

## Conclusions

The odds ratio of male mortality still remains high in the largest outbreak in a century caused by the pathogenic hCoV-SARS-CoV-2. Our data is not adjusted for the various factors that could drive these differences, nevertheless, there are growing concerns about how the virus could disproportionately impact the different genders^23,31^. Even though there is epidemiological data from pandemics, research and clinical practice do not reflect these distinctions. Ongoing research approaches should actively explore this area so that the knowledge derived can overcome this and prevent future pandemics. In addition we must collect disaggregated gender-relevant data that defines the various dimensions of this global epidemic. Moreover, aging studies in both sexes indicate diminished innate immune responses, and further analysis utilizing co-morbidities may provide the predilection for clinical worsening seen with this infection^13,32^. Taken together, registry data capturing socio-economic and co-morbidity characteristics and clinical protocols targeting the various components of the innate immune system should be explored to address the sex-specific differences in COVID infection.

## Data Availability

Studies chosen for these analyses were based on the availability of the sample size, and that of detailed patient demographics, including gender-associated incidence and mortality in peer- reviewed journals.

## Author Contributions

Conception and design: SPI, JE, and RC Manuscript writing: SPI, JE, and RC

Final approval of manuscript: SPI, JE, KA, PH, MD, AH, VS, CF, and RC

Accountable for all aspects of the work: SPI, JE, KA, PH, MD, AH, VS, CF, and RC

## Conflict of Interest

None to report

*Calculating the case-fatality ratio for a disease outbreak is difficult while the outbreak is still evolving. The true ratio cannot be determined until the outbreak is over when the total numbers of deaths and recoveries are known.

